# End-user perspectives on design and implementation of a novel SkinScan3D (SS3D) device for monitoring Kaposi Sarcoma in East Africa: a qualitative study

**DOI:** 10.64898/2026.08.12.26360308

**Authors:** Philippa Kadama Makanga, Harriet F. Adhiambo, Dorothy Mangale, Martha Nansereko, Jane Frances Nalubega, Roselyn Knight, Elvin Geng, Victor Mudhune, Elizabeth Bukusi, Fred Okuku, Aggrey Semeere, Thomas Odeny, Beryne Odeny

**Affiliations:** Infectious Diseases Institute, Makerere University, Kampala, Uganda; Washington University St Louis, USA; Kenya Medical Research Institute, Kisumu, Kenya; Uganda Cancer Institute, Kampala, Uganda

## Abstract

**Introduction:** SkinScan3D (SS3D) is a novel, artificial intelligence–enabled device that provides objective three-dimensional measurements for monitoring Kaposi Sarcoma (KS) lesions. Prior to launching a clinical trial of the device, we obtained end-user perspectives to guide device refinement.

**Methods:** Between April and May 2025, we conducted six focus group discussions and 28 in-depth interviews with patients, healthcare providers, and community representatives in Kenya and Uganda. Participants viewed a demonstration video and handled the SS3D prototype. Data were analyzed using hybrid deductive-inductive thematic analysis informed by the Health Information Technology Usability Evaluation Model and the Consolidated Framework for Implementation Research.

**Results:** Qualitative findings were synthesized into a conceptual framework for SS3D adoption with two interconnected themes: 1) experiences and context, and 2) device perceptions and implementation factors. Participants’ receptivity to the device was first shaped by experiences with medical technologies and the broader sociocultural context, including trust in providers, health beliefs, and gender preferences. After interacting with the prototype, participants viewed the SS3D as intuitive, accurate, and potentially capable of improving the objectivity and efficiency of KS lesion monitoring. They identified concerns related to safety, infection prevention, data security, affordability, maintenance, and workflow integration. Successful implementation was perceived to depend on device refinement, supportive organizational factors, including leadership engagement, provider training, maintenance capacity, and patient education to address misconceptions about the device. Participants proposed hardware, software, connectivity, and training refinements to support safe integration into routine clinical care.

**Conclusion:** End users demonstrated overall satisfaction and receptivity to the SS3D, given potential benefits for both patients and providers. We identified targeted refinements to optimize the device’s functionality and integration into the oncology environment to improve its fit with the local context.

## Introduction

Kaposi Sarcoma (KS) is the leading AIDS-defining cancer globally and a significant public health challenge, with the highest incidence and mortality in Africa.^1–4^ The greatest burden is in Eastern Africa, where KS is the third commonest cancer among men and a leading cause of cancer deaths in Uganda.^1^

Although treatment options for KS exist, monitoring lesions during treatment remains challenging. The current standard of care for monitoring KS lesions during treatment involves manually measuring the bi-directional diameters of the KS lesions.^5^ However, this approach is subjective, as measurements vary between providers, are time-consuming, especially in a busy clinic setting, and are frequently bypassed in routine practice. Yet, monitoring the size of KS lesions is important for patient staging, determining treatment response, and planning.^5^ Providers rely on the AIDS Clinical Trial Group (ACTG) staging to determine the prognosis of KS patients and plan treatment.^5^ Unreliable measurements can lead to inaccurate ACTG risk classification, prolonged therapy due to uncertainty of the treatment response, and unnecessary exposure to chemotherapy toxicity. Furthermore, the ACTG criteria perform poorly in persons with dark skin, compounding racial inequities in treatment-related adverse effects.^5^

More broadly, many medical devices reach routine clinical care yet are never adopted or are abandoned, largely because of prospective end users are inadequately involved during device development.^6, 7^ Eliciting end-user input before deployment is therefore an important step in ensuring that a new device is used in practice.

We sought to test the accuracy and reliability of a new device, the SkinScan3D (SS3D), in the Precision Imaging to Evaluate Kaposi Sarcoma study (PRIME-KS-Grant # U01CA292765).^8^ The PRIME KS study focuses on refining the function and evaluating the utility of the SS3D device. The SS3D is a low-cost device that uses liquid lens technology and artificial intelligence (AI). The SS3D provides three-dimensional (3D) images of KS lesions, including length, width, and height, enabling objective assessment of changes in lesion size and color. To refine the SS3D technology and usage protocols to inform future adoption, we conducted qualitative interviews with end users, including patients, providers, and community advocates in East Africa (EA). The objective of this work was to explore end-user experiences with implementing new health technologies, assess perceptions of the initial SS3D prototype, and identify potential implementation barriers, facilitators, and design modifications to enhance usability and contextual fit.

## Methods

### Study design and setting

Between April and May 2025, we conducted a qualitative study through Focus Group Discussions (FGDs) and in-depth interviews (IDs) to explore factors affecting the implementation of new technology and recommendations to refine the SS3D. We conducted the study in Jaramogi Oginga Odinga Teaching and Referral Hospital (JOOTRH), Homabay and Chulaimbo in Kisumu, Kenya, the Uganda Cancer Institute (UCI), and Infectious Diseases Institute (IDI) in Kampala, Uganda. JOOTRH is a government-funded teaching and referral hospital serving the general population in and around Kisumu County in western Kenya. The UCI is a semi-autonomous government-aided teaching and referral health facility that offers specialized oncology care for the whole of Uganda. Both JOOTRH and UCI provide treatment for patients with KS.

### Participants and sampling

We purposively sampled participants across three end-user groups — health providers (doctors, nurses, clinical officers, social workers, biomedical engineers, and regulatory officers), KS patients (identified via clinic records), and community representatives (from Community Advisory Boards and patient advocacy groups) — ensuring diversity by age, gender, and cadre. Recruitment was conducted by phone and in-person, and all participants provided written informed consent and were above 18 years.

### Data collection

We conducted six FGDs overall — one per participant group (health providers, patients, and community representatives) per country — and 28 IDIs across both countries (15 in Kenya, 13 in Uganda), using semi-structured guides with open-ended questions. During each session, participants watched a 5-minute demonstration video and physically handled the SS3D before sharing their perspectives. A trained interviewer and notetaker were present, and all sessions were audio-recorded and transcribed verbatim into English.

### Data analysis

We used a mixed deductive-inductive thematic analysis approach guided by the Health Information Technology Usability Evaluation Model (ITUEM) framework (Figure 1)^9^ and the Consolidated Framework for Implementation Research (CFIR) (figure 2).^10, 11^ ITUEM incorporates both subjective and objective measures to assess usability of information technology systems. ITUEM constructs applied deductively included satisfaction, efficiency, effectiveness, completeness and accuracy, information needs, flexibility, learnability, performance speed, and competency — applied to both prior device experiences and SS3D perceptions, on the premise that past technology encounters prime expectations along the same usability dimensions. Three CFIR domains were applied deductively: inner setting, intervention characteristics, and outer setting. We developed a codebook (BO, DM, HAF) guided by the frameworks and inductively added codes emerging from the data. Transcripts were uploaded to Dedoose and coded by eight coders (PKM, BO, DM, HAF, SO, MN, JFN, RK); thematic saturation was reached when no new insights emerged with additional interviews,^12^ and intercoder reliability was assessed with discrepancies resolved by consensus.

**Figure 1:**
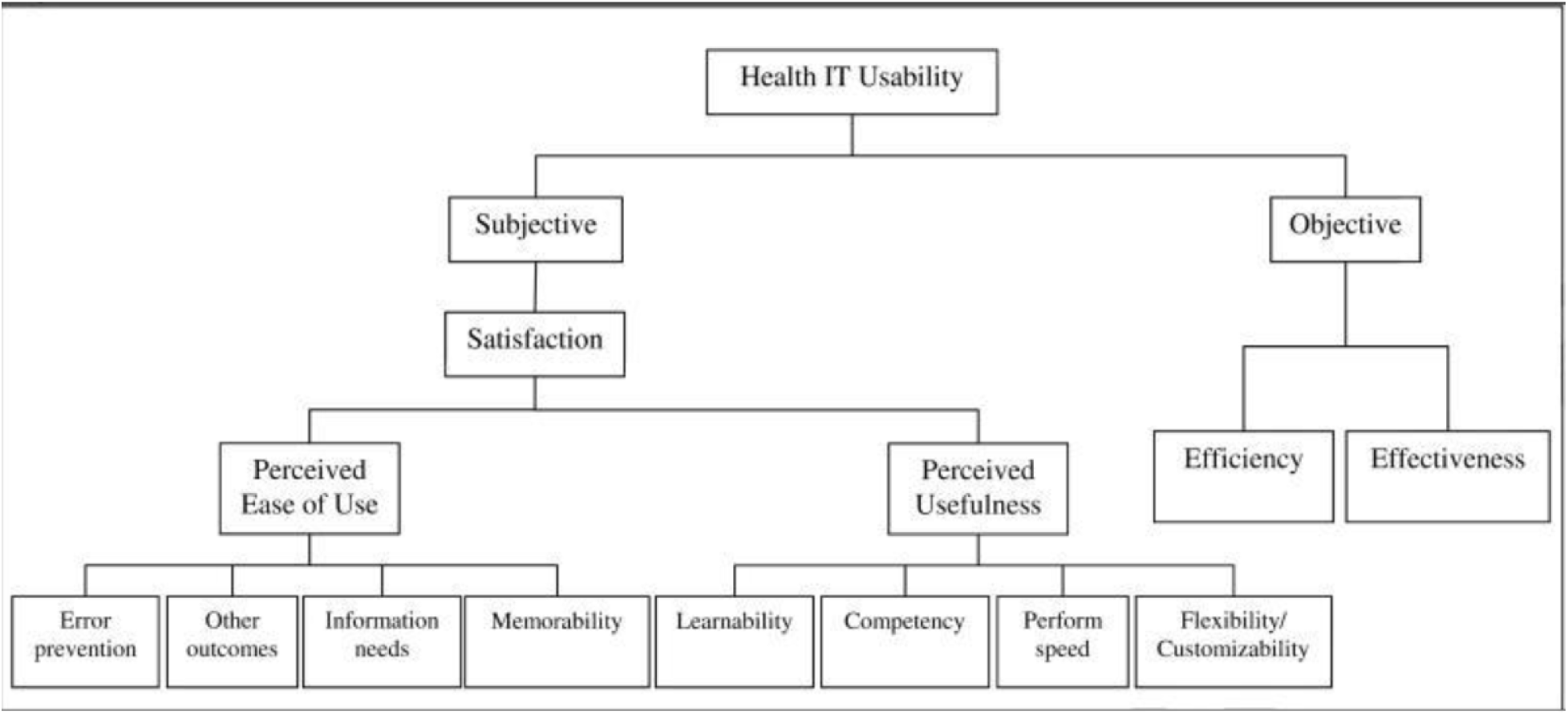
The Health IT Usability Evaluation Model. Adapted from Health Information Technology Usability Evaluation: Methods, Models, and Measures by P-Y Yen, 2010

**Figure 2:**
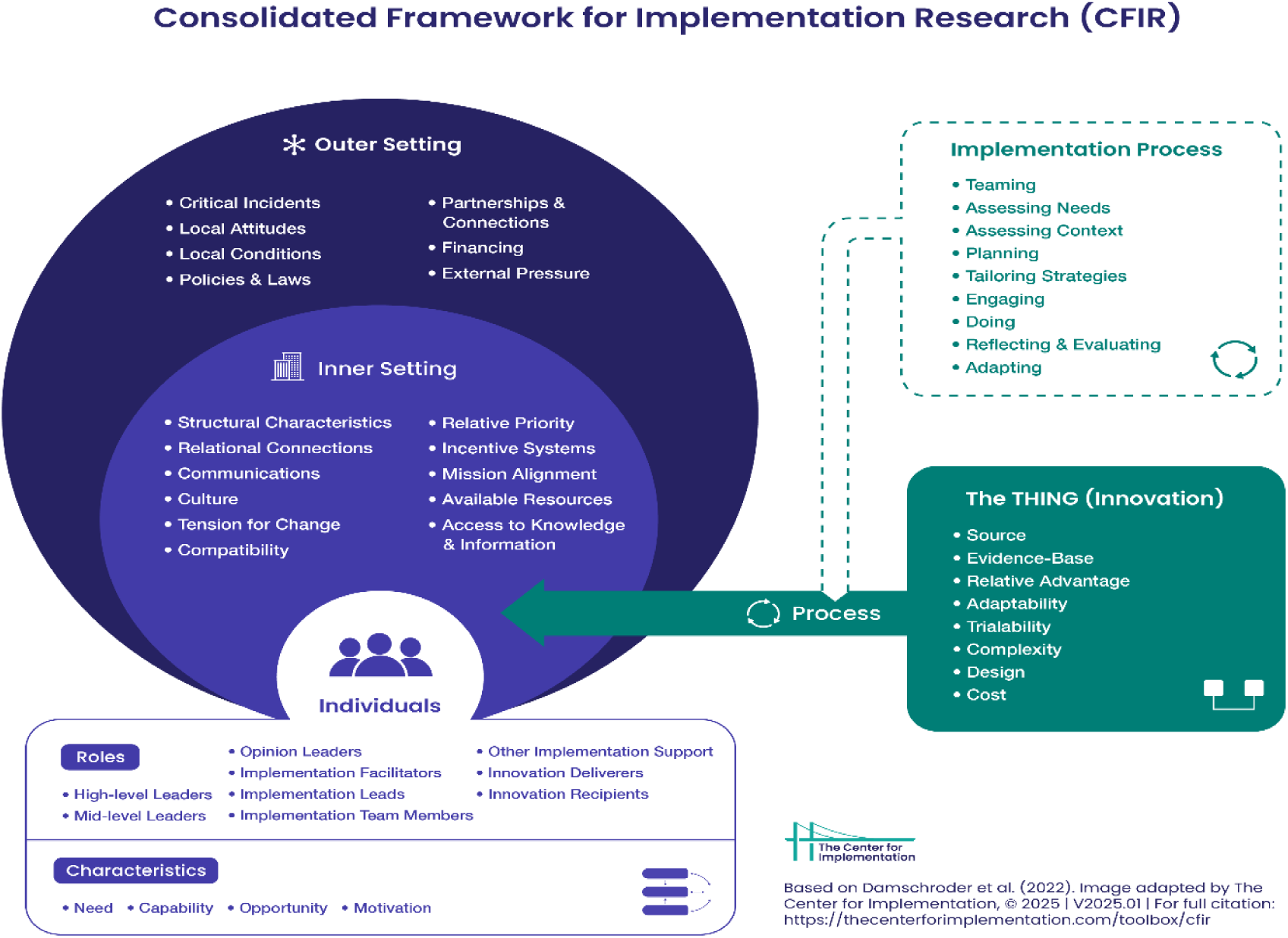
The Consolidated Framework for Implementation Research (CFIR). Adapted from The Center for Implementation, 2025

Methods are reported in alignment with the Consolidated Criteria for Reporting Qualitative Research (COREQ) guidelines^13^. The completed COREQ checklist is attached as supplementary material.

### Ethical considerations

All participants provided written informed consent before the interviews. We obtained ethical approval for the study from the JOOTRH Institutional Ethics Review Committee (JOOTRH-ISERC), reference number ISERC/JOOTRH/145/25 in Kenya, the IDI Research Ethics Committee (IDI-REC), reference number IDI-REC-2024-121 in Uganda, and the Washington University in St. Louis Institutional Review Board.

## Results

A total of 83 individuals participated in the FGDs (55 participants) and IDs (28 participants). Across all FGD participant groups, the majority in Kenya were male, while most in Uganda were female – Table 1.

**Table 1:** Demographic characteristics of FGD participants by country.

|  | **Kenya (N=32)** | **Uganda (N=23)** |
| --- | --- | --- |
| **Patients** |  |  |
| **Male (n, %)** | 8, 67% | 4, 57% |
| **Female (n, %)** | 4, 33% | 3, 43% |
| **Age in years (median, IQR)** | 37, (35-38) | 41, (36-43) |
| **Providers** |  |  |
| **Male (n, %)** | 5, 50% | 3, 37% |
| **Female (n, %)** | 5, 50% | 5, 63% |
| **Age in years (median, IQR)** | 44, (38-45) | 35, (31-39) |
| **Community members** |  |  |
| **Male (n, %)** | 4, 40% | 1, 12% |
| **Female (n, %)** | 6, 60% | 7, 88% |
| **Age in years (median, IQR)** | 40, (38-45) | 50, (39-55) |

The 28 ID participants are described in Table 2. Most Kenyan ID participants were male (64%), while the majority (54%) in Uganda were female. The median age of participants was 43 (IQR: 39-49) in Kenya and 39 (IQR: 32-44) in Uganda.

**Table 2:** Characteristics of In-depth interview participants by country.

|  | **Kenya, N= 15** | **Uganda, N=13** |
| --- | --- | --- |
| **Males (%)** | 10, 64% | 6, 46% |
| **Females (%)** | 5, 33% | 7, 54% |
| **Age in years (median, IQR)** | 43, (39-49) | 39, (32-44) |
| **Participant type (n, %)** |  |  |
| **Patients** | 2, 13% | 2, 15% |
| **Providers** | 8, 53% | 2, 15% |
| **Community representatives** | 5, 53% | 9, 69% |

### Overview of thematic findings

Through deductive and inductive coding, we identified two major themes. Theme 1, **Prior experiences and sociocultural context,** which describes how participants’ prior encounters with technology and sociocultural factors shaped their perceptions and receptivity to the SS3D. Theme 2, **Perceptions of the device and recommendations**, which describes participants’ perspectives and recommendations after handling the SS3D.

#### THEME 1: PRIOR EXPERIENCES AND SOCIOCULTURAL CONTEXT

Prior encounters with medical devices served as a frame of reference through which participants interpreted and evaluated the SS3D. Receptivity was modulated by the quality of these prior experiences with devices and, independently, by sociocultural factors operating at the individual and community level (**Table 3)**.

**Table 3.** Theme 1 – Prior experiences and sociocultural context: subthemes, codes, and quotes.

| Subtheme | Code | Illustrative quote |
| --- | --- | --- |
| **1A. Past experiences with devices** | **Satisfaction with past experience** | *"She thinks if you use that machine on her... she has gotten treatment. My aunt said, 'They have put a machine on me, I felt like the doctor checked me and I'm cured.'"* (Ugandan community representative) |
|  | **Completeness & accuracy** | *"If that machine does not give accurate results, then people may fear to use it or refuse to use it completely."* (Kenyan community representative) |
|  | **Information needs** | *"I think that what has made these machines normal is that a person has seen them for a long time. ... I think if they get enough teaching that person will find this machine usual."* (Ugandan community representative) |
|  | **Efficiency & cost** | *"I think it will help the doctor to work faster because the manual methods consume a lot of time "* (Kenyan KS patient) |
| **1B. Sociocultural factors** | **Societal beliefs & norms** | *"I've seen some patients who do not like their pictures being taken for any reason."* (Kenyan health provider) |
|  | **Gender concordance preferences** | *"... if she is female, then a female should work on her [i.e. use the device on her]. They feel comfortable..."* (Ugandan community representative) |

##### Subtheme 1A: Past experiences with devices

Below, we outline codes associated with this subtheme (supportive quotes for all codes are provided in Table 3).

###### Satisfaction with past device experience

Participants with positive past experiences were more receptive, expressed enthusiasm and openness toward the SS3D, and were satisfied with the use of devices during their care at health facilities. Some participants had negative experiences related to costs, accuracy, information needs and poor efficiency while using devices in the past.

###### Completeness and accuracy

Negative past experiences – particularly inaccurate results – led to mistrust of devices, which negatively affected their views and receptivity towards medical devices.

###### Information needs

Participants identified inadequate information about unfamiliar devices and related procedures as a barrier to receptivity of devices and a potential barrier to the use of the SS3D. Inadequate information led to fear, which reduced receptivity to devices, especially if the device is not usually used in patient care.

###### Efficiency and Cost

System delays and perceived financial exploitation in healthcare settings generated caution and a fear of the unknown. Participants cited long wait times and costs as potential barriers to adoption of the SS3D.

##### Subtheme 1B. Sociocultural factors

The sociocultural context shaped the participant’s perceptions and receptivity towards medical devices including SS3D. Codes and quotes are provided in Table 3.

**Societal beliefs and myths** may hinder the acceptance of medical devices as they can fuel fear and misconceptions of harms from devices.

###### Gender preferences

Particularly among adult women, gender concordance between the patient and device operator was an important consideration. Female patients preferred female operators when the device is used on them.

#### THEME 2: PERCEPTIONS OF THE DEVICE AND RECOMMENDATIONS

After handling and interacting with the SS3D prototype, participant feedback was grouped into four subthemes: Usability perceptions, Satisfaction and safety, Device attributes, and Implementation environment. (**Table 4)**.

**Table 4:** Theme 2 — Perceptions of the device.

| **Subtheme** | **Code** | **Illustrative quote** |
| --- | --- | --- |
| **2A. Usability perceptions** | **Learnability** | *"Yes, and it's also easier to understand. You just only need on-the-job training, so that even if I'm trained to use it, I can train my colleague, and the colleague can train the other." (Kenyan health provider)* |
|  |  | *"I even see some of the most illiterate people operating iPhones. Someone who has trained as a healthcare worker cannot fail to use that equipment." (Kenyan health provider)* |
|  | **Completeness & accuracy** | *"It will avoid a lot of guesswork... you have the initial measurement, you have the current measurement and you do the comparison. It might also avoid inter-reader biases." (Kenyan health provider)* |
|  |  | *"As health providers, it will help to display whether the lesions have cleared or not." (Ugandan KS Patient)* |
|  | **Efficiency & performance speed** | *"Basically, the workflow will be now easier, well-organized and timely."*<br>*"I think it will help the doctor to work faster because the manual methods consume a lot of time " (Kenya Health Provider)*<br>*"They will like the technology because it will reduce on the time the patient will spend in the doctor's room. It speeds up the process, ... it will give opportunity for many patients to see the doctor." (Ugandan Community representative)*<br>*"At times they look at it as if you are wasting their time. The machine takes time to boot. He would rather use a ruler." (Ugandan health provider)* |
| **2B. Satisfaction & safety** | **Provider & patient satisfaction** | *"I think the most good thing there is, is the advantage to the patient knowing that either you're improving or actually you're deteriorating and either the regimen that you're on — treatment is working or it's not working." (Ugandan health provider)* |
|  |  | *"personally, I want to see whether my scar is increasing in size or reducing in size which right now I'm not certain about, but with the presence of this machine, it would be of great help" (Ugandan KS patient)* |
|  | **Infection prevention** | *"When you use this on a patient, is it cleaned before the next patient?" (Kenyan health provider)* |
|  | **Psychological benefits** | *"It will give us confidence that we shall get cured since the supervision has increased. Because you have not just observed my condition, but tested me using this machine. So I expect that I will be getting better." (Ugandan KS patient)* |
| **2C. Device attributes** | **Flexibility/ customizability** | *"But I love the fact that it's portable. We don't need to install it in a room. We don't need to build a special room to install it... We just need to charge and clean the probes for reuse." (Kenyan Health Provider)* |
|  |  | *"The fact that this thing can be charged and work even during power outages means there should be no disruption of care. Even during those times when you don't have electricity. " (Kenyan Health Provider)* |
|  |  | *"Everything would fit in a very small box. I would just be worried that when it's very portable then theft is easier." (Kenyan Health Provider)* |
|  | **Hardware design** | *"The plastic material...It's light and the beauty is it doesn't break, ... if you go for glass and something of the sort then issues of breakage arise." (Kenyan Health Provider)*<br>*"This cable the way it is (touches the cable), it can easily attract germs. If it could have been made to be something smooth, it would be easy to clean." (Kenyan health provider)*<br>*"...I think it would be best if we have more cups....?" (Kenyan Health Provider)* |
|  | **Software & data security** | *"...what I might like is first of all the graphics; it's touchscreen like you demonstrated. Most of the things are touch. There will be less buttons to press ... " (Ugandan Health Provider)*<br>*"... there might be some data privacy concerns. Handling high precision images for patients, they require secure data management to prevent breaches." (Ugandan health provider)*<br>*"I think also for the purposes of confidentiality we need some password for this gadget for you to get in and also log out when done" (Kenyan Health Provider)* |
|  | **Device cost** | *"It should be cost effective, affordable, so that when it has broken down, we can afford to purchase another one or we can purchase more pieces." (Kenyan health provider)*<br>*"What is the maximum cost that as a hospital we will be willing to incur to get the additional benefit that the gadget brings in terms of monitoring the lesion size in the light of alternative methods that have always been used?" (Kenyan Health Provider)* |
| **2D. Implementati on environment** | **Leadership engagement & inner setting** | *"For leadership, you know, for it to actually trickle into where you are, they must accept the device, and they must understand the purpose of why it's going to be used." (Kenyan health provider)*<br>*"... the machine itself, breaking down..., are we going to have the bioengineers taught about the machine, how to prepare it and everything? I mean, machines always break down, so we need at least someone who is going to do it, giving it service and preparing and everything." (Ugandan Health Provider)*<br>*"...As we know, not everybody is going to embrace this. Some people are going to resist and stick to their own methods of doing things. As we know, not everybody accepts change." (Kenyan Health Provider)* |
|  | **Patient health literacy & education** | *"... they come with different beliefs and perceptions, so you have to handle them as they present to you, you need to talk to them, educate them as well on the new technology that is coming up. You need to take time to teach." (Kenyan health provider)* |

##### Subtheme 2A: Usability perceptions

Three ITUEM constructs – learnability, completeness and accuracy, and efficiency/performance speed – captured participants’ assessments of how the SS3D would perform in routine clinical use. *Supportive quotes for all codes are provided in Table 4*.

###### Learnability

In both countries, all participant groups perceived the SS3D as learnable and feasible to operate with appropriate training. The touchscreen interface was considered familiar and intuitive, drawing comparisons to smartphones. No participant considered specialist-level technical skills a requirement to operate the SS3D. While SS3D was perceived as easy to learn, adequate training was universally expected.

###### Completeness and accuracy

The most consistent finding across all groups was the anticipated clinical value of objective, reproducible lesion measurement. Participants recognized inter-provider variability in manual measurement as a limitation the SS3D would address. Patients also valued the transparency of objective measurement, reporting that knowing whether their lesions were improving or worsening reduced anxiety and strengthened treatment engagement.

###### Efficiency and performance Speed

Participants generally anticipated that the SS3D would improve clinical efficiency. There was an expectation of a more efficient clinic workflow, improved organization and reduced clinic time for patients. Nevertheless, a contrasting negative perception regarding boot-up time was a concern for providers accustomed to the convenience of manual measurement. Providers made recommendations to minimize device startup time.

##### Subtheme 2B: Satisfaction and Safety

Overall satisfaction was high across participant groups but was modulated by a primary safety concern from providers about the absence of a clear infection prevention protocol for the probe cups.

###### Provider and patient satisfaction

Providers were satisfied with the objectivity of the SS3D to help them see any changes in KS lesions that would guide them on future patient management. Patients were satisfied to have objectivity in assessment of lesion status, which reduced anxiety regarding whether the treatment was working or not.

###### Infection prevention

The absence of a clear infection prevention protocol for the cup, was concerning. Recommended solutions included per-patient disinfection protocols, disposable cup covers, and patient-specific cups.

###### Psychological benefits

For patients, the psychological benefit of objective measurement—knowing whether lesions were improving reduced anxiety about treatment efficacy—was important.

##### Subtheme 2C: Device attributes

Participants assessed the SS3D’s physical and digital attributes across four codes (Table 4): flexibility/customizability, hardware design, software and data security, and device cost. These mapped onto the CFIR “Intervention characteristics” domain and the ITUEM domains.

###### Flexibility/customizability

Participants valued the SS3D’s portability and battery-powered operation, particularly in the context of frequent power outages. The 36-hour battery life was viewed as a facilitator for uninterrupted use. However, portability raised a theft concern and the need for safety measures to be in place. Regarding customizability, participants were also concerned about the size of the cup not being able to cover varying lesion sizes, and they recommended provision of cups of different sizes.

###### Hardware design

The polycarbonate construction was lightweight, durable (not breakable), and potentially easy to clean. There were concerns including the cable surface (perceived as prone to contamination), probe length, and the durability of the device in busy environments.

###### Software and data security

Participants appreciated the touchscreen interface and real-time display. There were concerns around data security as unauthorized access to high-resolution patient images. Recommended additions included password-protected logins for all providers. Other desired enhancements included solar charging capability, alternative power sources for rural facilities, and wireless/Bluetooth probe connectivity.

###### Device cost

Participants recognized that the SS3D would need to be affordable at initial purchase and for ongoing maintenance and replacement. Facility administrators questioned incremental benefit relative to cost compared to manual method for KS lesion measurement.

##### Subtheme 2D: Implementation Environment

Three codes addressed the broader context required for SS3D adoption beyond perceptions of the device itself.

###### Leadership engagement and inner setting

Health facility leadership engagement was identified as essential for sustained adoption. Providers emphasized that without leadership buy-in and maintenance infrastructure — including trained maintenance staff at each site — adoption would be challenging after the research period. Resistance to change from some providers was also noted as a challenge to prepare for.

###### Patient health literacy and education

A distinct finding was the widespread conflation of the SS3D with a treatment device. Participants consistently perceived the device as therapeutic rather than monitoring in function, even after explanation. Patient education was recommended to address misconceptions about the SS3D as treatment rather than a monitoring device, and to address health literacy barriers. In Table 4, we summarize the major themes, subthemes, codes, and quotes.

##### Key Recommendations

Participants generated both device design-related and implementation recommendations (Table 5) to enhance the device’s usability and adoption.

**Table 5:**
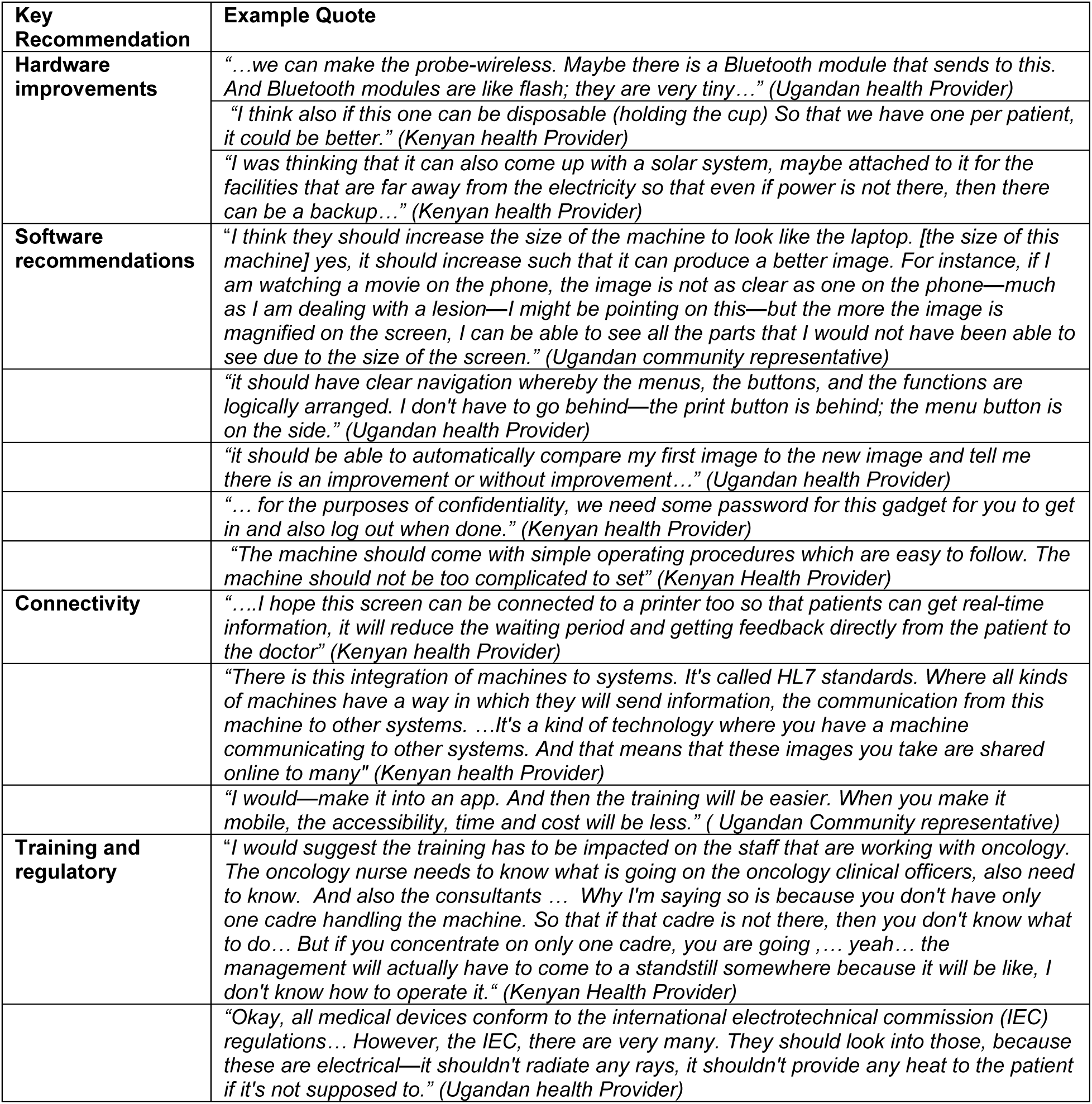
Key recommendations to optimize the SS3D and facilitate its implementation.

###### Hardware

Participants provided recommendations to prevent infection, including future development of a wireless (Bluetooth or laser) probe, the use of multiple or disposable (single-use cups), disinfection protocols, and using a shorter, smoother cable to avoid dirt accumulation in the cable’s ridges. There were suggestions to use alternative power sources, such as solar energy, especially in rural settings.

###### Software

Enhancements to improve device image quality, security, and functionality were suggested, including a bigger screen size to enhance image quality, password-protected logins for improved data security and privacy, AI-assisted automatic image comparison between visits for easy comparison of changes in the KS lesions between visits, and clear on-screen navigation menus to enhance the user experience. A user troubleshooting manual was also recommended.

###### Connectivity

Participants were keen to have the SS3D interoperable with other devices and hospital information systems to facilitate communication with patients and among various sections of the health facility. Suggestions included connecting the SS3D to a printer for real-time results or making it Health Level Seven (HL7) compatible to allow communication with other hospital systems. Some health providers suggested the development of a Mobile Application to enhance access and scale up this technology.

###### Training, regulatory measures & leadership engagement

Participants recommended adherence to industry standards, training providers, patient education, and addressing information needs. Initial training of multiple cadres (with comprehensive training materials) would ensure that staff are available to operate the device without interruption during staff turnovers. In addition, well-trained maintenance staff and user manuals are needed for smooth and sustained device function. Compliance with industry safety, along with leadership engagement, was suggested to facilitate easy integration and rollout. Patient education was recommended to eliminate misconceptions around the device’s therapeutic capability and emphasize its monitoring function.

## Discussion

This study assessed end-user perspectives on the novel SS3D device for KS lesion monitoring in Kenya and Uganda and identified refinement priorities to optimize the device before its deployment. Many medical devices fail to achieve routine clinical use because end users are not adequately involved during development, thus early engagement in needed to optimize device usability and adoption.^6,7^ Usability and adoption of technology are shaped by multiple factors,^6^ including inadequate end-user input during development.^7^ This study is essential to ensure the SS3D is suitable for clinical practice before its use in a clinical trial. Our findings –organized around an emerging conceptual framework for SS3D adoption (Figure 3) – highlight the interplay of factors most relevant to the local context. The framework draws together three interconnected domains, including experiences and context, perceptions of the device, barriers and recommendations that will shape SS3D prototype iteration.

**Figure 3:**
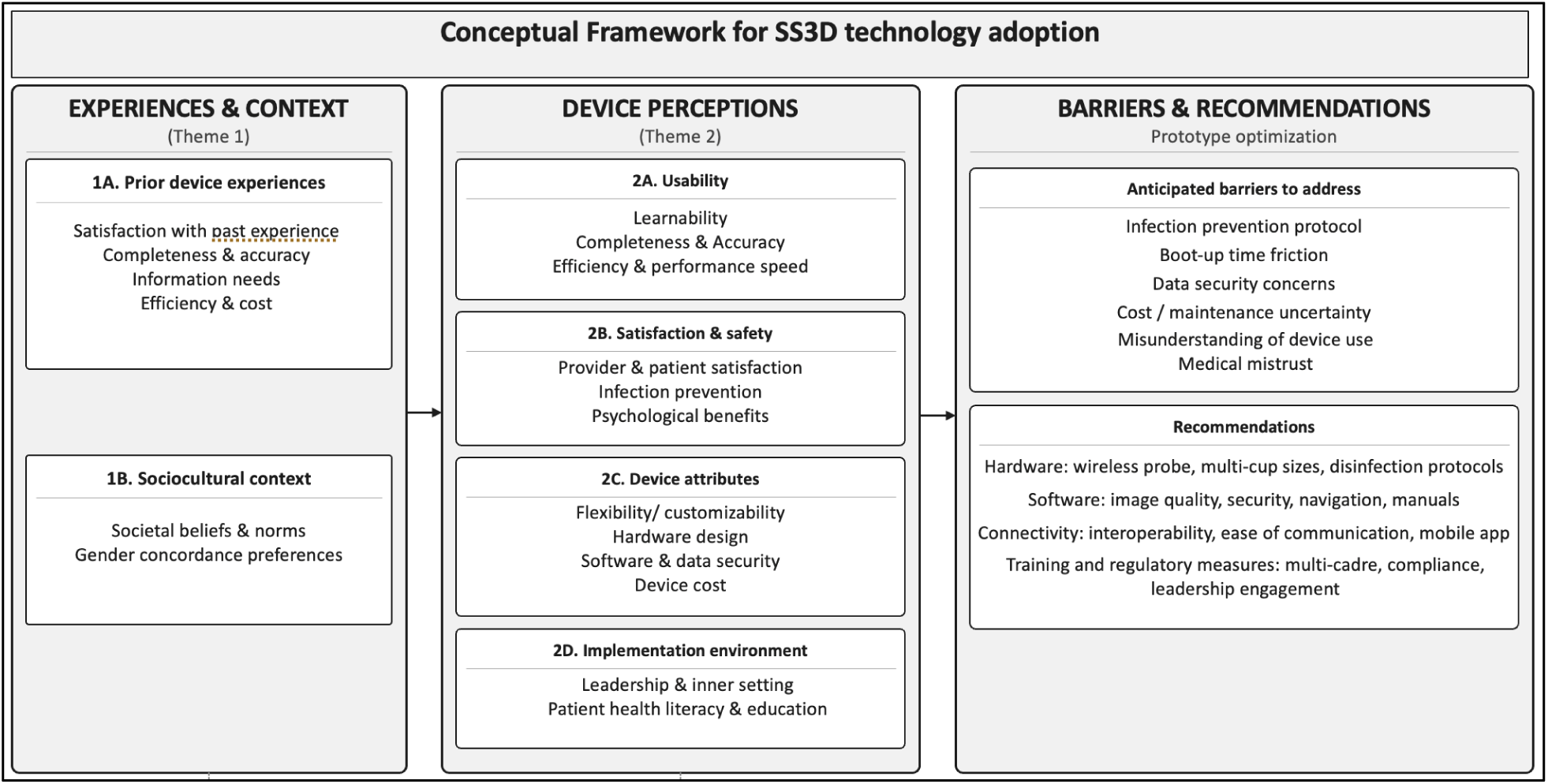
Conceptual framework of SS3D adoption: bringing together themes, subthemes and codes

### Experiences and context

As anticipated, prior technology experiences shaped participants’ perceptions of and receptivity to the SS3D in both positive and negative ways. Positive experiences with prior devices led to openness and support for SS3D, while negative experiences — including fear of unfamiliar technology, inaccurate results, and financial exploitation — emerged as potential adoption barriers. This is consistent with evidence that prior health system and technology interactions directly inform receptivity to new devices, underscoring the need to address the experiences of the target population prior to technology deployment.^14, 15^

Sociocultural factors and misconceptions around treatment capability of the device and harms emerged as determinants of receptivity to SS3D and its adoption. Gender concordance preferences between patients and providers were also important determinants of device receptivity.^16^ Participants highlighted beliefs and myths about electronic devices — most strikingly, the misconception that SS3D is for treatment rather than monitoring. Sociocultural factors and technology misconceptions are documented barriers across sub-Saharan Africa, particularly where health literacy is constrained by limited formal education and low prior exposure to diagnostic devices.^17, 18^ Cultural- and gender-sensitive tools, operating protocols, and patient education – to clarify the device’s function – are essential to mitigate unwarranted fears and expectations while emphasizing the device’s benefits.^19^

### Perceptions of device

Device accuracy, learnability and user-friendliness were important facilitators of adoption. Overall satisfaction with the device was high as participants perceived it as intuitive and trainable. SS3D’s accurate and reproducible measurement capability was valued as a direct improvement over current manual assessment. These observations align with systematic review evidence on ease of use and technical barriers as determinants for adoption of digital health technologies.^20^ Additionally, the patient-doctor relationship was affected by the technology. As seen in another study that assessed patient concerns with AI, medical trust—shown by the participants’ willingness to engage with SS3D if introduced by a trusted provider—was a gateway to device acceptance and adoption.^21^ Psychological benefits were reported from knowing lesion status, especially when treatment response is slow. However, infection prevention, workflow disruptions, and data security were key areas to prioritize during device optimization. Addressing these areas, particularly through provision of multiple sterilizable cups, staff training and workflow integration, software refinement, and ensuring password-protected access, is essential for device adoption as documented in other studies.^22, 23^

Cost and the health facility environment are important determinants of SS3D adoption. Initial purchase and maintenance costs could undermine the device’s incremental benefits. Use of the SS3D in this setting is more likely if it is affordable to purchase and maintain, and health facility leadership is engaged and supportive.^24^ Planning for future SS3D rollout will incorporate leadership engagement strategies as a core component. Cost-effectiveness evaluations comparing SS3D to conventional manual measurement are a priority for future research to support organizational buy-in.

### Barriers and recommendations

The findings highlight key barriers such as infection prevention, boot-up time and slow device speed, data security, purchase and maintenance cost, misconception of device use, and medical mistrust. Suggested recommendations were grouped into the hardware, software, connectivity, training and regulatory domains on the SS3D adoption framework. There were regulatory and ethical recommendations to boost data security, privacy, and storage, which align with systematic review evidence on device functionality and data management as barriers to the adoption of point-of-care devices.^25^Together, these recommendations will guide our engineering and clinical teams in refining the next SS3D prototype.

### Limitations

Participants had only brief prototype exposure rather than sustained clinical use, and perceptions may shift with real-world integration. Purposive sampling may over-represent participants already engaged with clinic-based research, potentially introducing positive receptivity bias. Findings were generated at referral-level oncology facilities, and adoption barriers may differ in lower-resource, lower-volume settings where KS care infrastructure is less developed.

### Strengths

The multi-site, multi-stakeholder qualitative design across two countries provides rich qualitative data and a strong foundation for SS3D refinement. The analysis was robust as it was guided by established frameworks and leveraged the intercoder reliability across coders. All interviews were conducted in English and the native languages, as applicable, which expanded the depth of expression for participants and diversity of insights.

## Conclusion

End users were receptive to the SS3D and willing to use it in clinical practice, with satisfaction driven by its ease of use, potential accuracy, and perceived utility for KS lesion monitoring. The refinements identified here will inform the next prototype before deployment in the PRIME-KS trial, and the lessons drawn offer a practical foundation for researchers introducing novel monitoring devices in resource-limited oncology settings across East Africa.

## Data Availability

All data produced in the present study are available upon reasonable request to the authors

## Acknowledgments

We appreciate the study teams at JOOTRH, IDI and UCI for their efforts during the implementation of this work. The authors acknowledge the patients, healthcare providers, and community members who participate in this study. We thank Susan Krown, MD, for her advisory role on staging and response criteria for KS. Joseph Carson’s contributions were supported in part by U.S. National Science Foundation Award 2242812 and U.S. National Institutes of Health Award P20GM103499. Philippa Kadama Makanga was supported with protected time during the final stages of write-up of this manuscript by the Fogarty International Center of the National Institutes of Health under Award Number D43TW009343 and the University of California Global Health Institute.

## Funding and Author Declarations

This study is funded by the National Cancer Institute (NCI), U.S. National Institutes of Health, grant number U01CA292765. The SkinScan3D devices are manufactured and provided by Pensievision. The funders had no role in the design of the study, the writing of this protocol, or the decision to submit for publication. The funders had no role in data collection, analysis, interpretation, or the decision to publish trial results.

The study protocol was registered at https://ClinicalTrials.gov NCT06898203, registered 27 March 2025. Pan African Clinical Trials Registry PACTR202603523439856.

## Notes

**Acknowledgments of research support**: This study is funded by the National Cancer Institute (NCI), U.S. National Institutes of Health, grant number U01CA292765.

### Competing Interest Statement

The authors have declared no competing interest.

### Author Declarations

JOOTRH Institutional Ethics Review Committee of Jaramogi Oginga Odinga Teaching and Referral Hospital gave ethical approval for this work. IDI Research Ethics Committee of Infectious Diseases Institute, Makerere University gave ethical approval for this work. Institutional Review Board of Washington University in St. Louis gave ethical approval for this work.

